# Trends and Predictors of Measles Immunisation Inequalities Among Children Aged 12–23 Months in the Philippines, 2008–2022

**DOI:** 10.64898/2025.12.01.25341428

**Authors:** John Carlo Lorenzo, Md Irteja Islam, Carla Ante-Orozco, Erin Mathieu

## Abstract

**Introduction:** The Philippines continues to experience measles outbreaks with declining coverage of the first dose of the measles vaccine (MCV1). Despite various interventions and policies to improve MCV1 coverage, inequalities persist, influenced by various socio-demographic factors. This study aims to identify predictors and examine trends in socio-economic inequalities in MCV1 receipt.

**Methods:** Using the four most recent Philippine DHS datasets (2008, 2013, 2017, and 2022), we identified predictors of MCV1 receipt through bivariate and multivariable regression models. We examined trends in socio-economic inequalities in MCV1 using the Slope Index of Inequality (SII) and Relative Index of Inequality (RII).

**Results:** Our study included 5,584 children aged 12–23 months. Overall MCV1 coverage declined from 84.7% (2008) to 80.8% (2022). Significant predictors of MCV1 receipt included household wealth quintile, maternal education, parity, ANC visits, distance to health facility, and maternal smoking status. Socio-economic inequalities in MCV1 decreased from 2008 (SII=0.27; RII=1.41) to 2013 (SII=0.23; RII=1.33), stagnated in 2017 (SII=0.23; RII=1.35), and increased markedly by 2022 (SII=0.31; RII=1.52). Health system constraints including inadequate vaccine supply, suboptimal cold chain infrastructure, and limited trained immunisation staff disproportionately affected disadvantaged communities. The 2017 Dengvaxia controversy has eroded public trust in vaccines, while recurrent measles outbreaks and the COVID-19 pandemic further disrupted vaccine access.

**Conclusion:** Sustaining and monitoring equity-focused interventions, strengthening primary healthcare through Universal Health Coverage, and rebuilding community trust and engagement are essential to ensure measles vaccines remain available, accessible, and acceptable to disadvantaged communities experiencing measles outbreaks and immunisation inequalities.

## Introduction

Measles remains an important cause of childhood morbidity and mortality. It is a highly infectious vaccine preventable disease (VPD), requiring 95% coverage for measles vaccines to prevent outbreaks [1]. Preliminary data from the World Health Organisation (WHO) and UNICEF shows that in 2024, an estimated 395,000 measles cases were reported globally, with surges of infections in many regions [2]. Measles can be prevented with life-saving vaccines; however, the COVID-19 pandemic stalled the global coverage of measles, leaving children vulnerable to infections, contributing to recent measles outbreaks in many countries [3].

In the Philippines, where measles remains endemic, recurrent outbreaks were reported prior to the COVID-19 pandemic, with over 48,000 cases reported in 2019 [4]. This trend has declined in 2021 to 2022 due to measles surveillance setbacks [3], COVID-19 infection control measures, and temporary closure of schools and crowded areas which reduced the transmission of many respiratory infections [5]. According to the Department of Health (DOH) Philippines, nearly 900 children have died due to measles from 2018 to 2023 [6], and subnational surges of measles infections are picking up again in the country since 2023, particularly in the Southern regions. The Bangsamoro Region, one of the poorest and inaccessible regions in the country, experienced the longest measles outbreak requiring the vaccination of more than 1.2 million children as a supplementary response in 2024 [7]. As of mid-2025, the Philippines recorded over 2,068 measles cases from January to May, marking a significant increase compared to the same reporting period in 2024 [8].

Measles immunisation in the Philippines is under the Expanded Programme on Immunisation (EPI) which has been established since 1976, with the inclusion of anti-measles vaccines in 1982 [9]. Since then, the EPI has set the target to eliminate measles in the country with an over-all goal of reducing childhood morbidity and mortality from VPDs. Measles vaccines are provided for free in public hospitals and public health centres for children under-five years as mandated in the Republic Act no. 10152 of 2011 [10]. Measles coverage for both the first (MCV1) and second dose (MCV2) has not achieved the 95% target since 2000, with 2024 coverage of 80% and 71% for MCV1 and MCV2, respectively. Despite efforts to increase measles immunisation coverage in the country through strategies such as Reaching Every Barangay (REB), Supplemental Immunisation Activities (SIA) for measles vaccines, and sustained routine surveillance of measles and other VPDs, declining coverage highlights significant gaps in reaching children left behind by national and subnational strategies.

Various challenges contribute to the consistently low measles immunisation coverage. These include the readiness of the health system to operationalise immunisation programmes, which affects vaccine availability and quality [11], as well as contextual factors, attitudes, and personal beliefs [4] that influence the demand for vaccination. Vulnerable populations identified by the Immunisation Agenda 2030 (IA2030) [12] as being left behind by immunisation programmes are expected to bear the burden of both immunisation service delivery and uptake challenges. For instance, the marginalised and poor communities would be excluded from the vaccine benefits due to poorly designed and suboptimal immunisation programmes [13] while also being vulnerable to misinformation of measles vaccines highlighted by the level of education [14].

A substantial body of evidence has investigated wealth inequalities in childhood immunisation, however, much of this research focused at the regional or global level [15], [16], leaving local context and specific characteristics associated with these inequalities underexplored. The outcomes of our recent study [17] recommended examining the trends of wealth inequalities in immunisation over time to better understand how they evolve. Additionally, few studies have investigated measles immunisation as a standalone outcome, underutilising its potential as both an early warning indicator for outbreaks of other VPDs and as a marker of challenges in immunisation programme implementation [12]. As highlighted by the IA2030 framework and Mulholland’s study on child survival [12], [13], monitoring both coverage and equity in measles vaccination programmes is critical for identifying gaps and informing strategies to address them. This study assesses trends in MCV1 coverage across socio-demographic factors, identifies predictors of MCV1 receipt, and examines the trends in socio-economic inequalities in measles immunisation in the Philippines.

## Methods

### Data source

We utilised data from four recent Demographic Health Surveys (DHS) of the Philippines conducted in 2008, 2013, 2017, and 2022, which obtained comparable nationally representative samples of children aged 12–23 months [18]. Access to DHS metadata was sought from the Philippine Statistics Authority (PSA). The DHS is an extensive, nationally representative, and large-sample size survey that covers multiple indicators including vaccination status of children under-five years from more than 90 low-middle-income countries (LMICs) [18], [19]. The DHS utilises two-stage cluster sampling to select samples. The first stage determines the primary sampling units (PSU) using probability proportional to size from a national master sample frame. During the second stage, a sample of households are selected from the complete list of households in each selected PSU [18], [19].

### Study population

This study included children aged 12-23 months with data on the first dose of measles containing vaccine (MCV1) and complete data on selected explanatory variables. This population provides a reliable measure of measles vaccination coverage and aligns with WHO and DHS standard practices for immunization assessment [20], [21]. **Figure 1** shows the exclusion and inclusion criteria in the selection of study samples.

**Figure 1:**
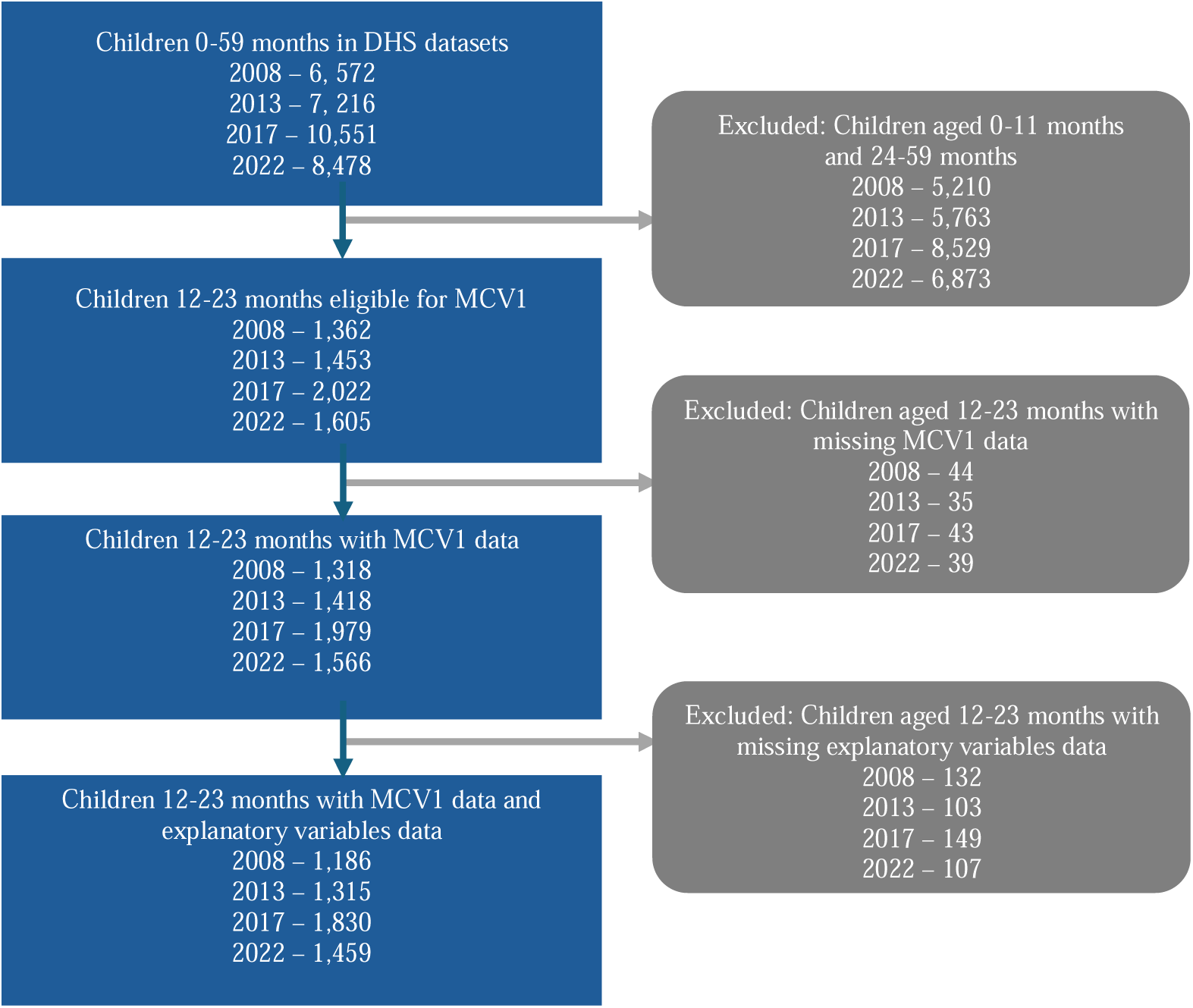
Selection of study participants

### Measures

Our main outcome variable was the receipt of MCV1 among children aged 12–23 months. If physical vaccination cards were not available, interviewers of DHS relied on self-reporting of mothers to ascertain immunisation status. Based on published literature, we selected nine explanatory variables from the DHS datasets to assess potential predictors of receiving MCV1 immunisation. This includes household wealth quintile (poorest, second, middle, fourth, richest), sex of the child (male, female), maternal age (15-19, 20-29, 30-39, 40-49), maternal education (no education, primary, secondary, higher), place of residence (urban, rural), parity (one to two, more than two), antenatal care (ANC) visits (fewer than four, four or more), distance to health facility (big problem, not a big problem), and maternal smoking status (yes, no). We used the smoking status of mothers to represent parental health behaviour and experiences as discussed in the literature [22]. **Supplementary Table A** summarises the predictors and their coding and definition used in this current analysis.

### Statistical analysis

We used the *svy* command in Stata (Stata19.0) to account for the complexity of DHS survey design including adjustment for clustering, stratification, and sample weights [23]. For each DHS year, we have calculated the MCV1 coverage and the 95% confidence interval (CI) by socio-demographic characteristics to assess how MCV1 coverage was changing according to socio-demographic dimensions from different survey years. We conducted bivariate analysis and multivariable logistic regression in the pooled samples to identify significant predictors of receiving MCV1 dose, with only those statistically significant (P<0.05) from the univariate analysis were included in the multivariable logistics regression model.

To examine trends in inequalities in measles vaccination coverage by socio-economic strata, we calculated for both absolute inequalities and relative inequalities per DHS survey year. We used absolute difference (AD) and the Slope Index of Inequalities (SII) to present absolute inequalities, while relative difference (RD) and Relative Index of Inequalities (RII) were used to present relative inequalities in MCV1 receipt according to socio-economic status. The ADs were calculated by subtracting the coverage of measles immunisation in the poorest quintile (Q1) from each of the remaining wealth quintiles (Q2, Q3, Q4, and Q5). Similarly, RDs were calculated by dividing each wealth quintile (Q2, Q3, Q4, and Q5) by the poorest quintile (Q1) [24], [25].

The SII and RII presents the absolute and relative differences in measles immunisation between two extreme categories, in our case those children whose wealth scores are grouped in the poorest quintile (Q1) and those children whose wealth scores are grouped in the richest quintile (Q5). We ranked children aged 12-23 months using the wealth index factor score available in the DHS datasets and based on their position in the cumulative distribution we have allotted a relative rank for each child. For each quintile, children were then analysed based on the range and midpoint within the current distribution. Using a generalised linear model, we then regressed the MCV1 status against the midpoint with a logit link to obtain the predicted values for Q1 and Q5. The SII and RII were then calculated using the difference and ratio, respectively, of predicted values of Q1 and Q5 [25].

For AD and SII, a positive value indicates that the concentration of inequality is towards higher quintiles compared to Q1, while a negative value indicates that the inequality is among the poorest. Noting that the higher magnitudes indicate higher inequalities and the inequalities measured by these two measures are significant when the 95% CI does not cross the zero value. On the other hand, RD and RII can only produce a positive value with results that are >1 indicating higher concentration of inequalities towards the higher quintiles, while the resulting RII value of <1 indicates concentration of inequalities among the poorest quintile, and a resulting RII value of 1 indicates that there was no inequality observed among the poorest and richest wealth quintile groups. Noting that the inequalities measured by these two measures are significant when the 95% CI does not cross the value of 1 [25].

## Results

Once we adjusted for sampling design and weighting variables, our studies included a total of 5,584 children aged 12-23 months from the recent four DHS conducted In the Philippines. Sample sizes per DHS year were: 2008 = 1,163; 2013 = 1,296; 2017 = 1,790; and 2022 = 1,335.

Our study samples have more children belonging to the poorest quintile (Q1). Majority of mothers of children in the study were aged 20 to 29 years (2008 = 51.2%; 2013 = 49.5%, 2017 = 49.0%; and 2022 = 44.8%) and had at least had some secondary schooling (2008 = 48.5%; 2013 = 50.5%, 2017 = 54.5%; and 2022 = 48.1%). Across the surveys, there is an equal distribution of sex of the child, place of residence and parity. While there has been a significant increase in the number of mothers having higher education, the number of women who have completed four or more antenatal care visits, and a greater number of households indicating that distance to the health facilities is not a big problem, highlighting efforts to bring healthcare closer to the communities. Lastly, in all DHS years included, we found a consistently low proportion of mothers who smoke (2008 = 4.8%%; 2013 = 6.4%, 2017 = 5.6%; and 2022 = 3.8%). These trends are summarised in **Table 1**.

**Table 1:** Trend in socio-demographic characteristics of children aged 12–23 months (2008, 2013, 2017, and 2022)

| Dimensions | 2008 (95% CI) N = 1163 | 2013 (95% CI) N = 1296 | 2017 (95% CI) N = 1790 | 2022 (95% CI) N = 1335 |
| --- | --- | --- | --- | --- |
| **Wealth quintile** |  |  |  |  |
| Poorest | 24.37 (21.35, 27.67) | 24.93 (22.40, 27.63) | 26.59 (23.90, 29.46) | 29.54 (25.87, 33.50) |
| Second | 22.26 (19.76, 24.97) | 20.61 (18.38, 23.03) | 20.93 (18.34, 23.78) | 17.66 (15.14, 20.51) |
| Middle | 20.48 (17.94, 23.28) | 23.42 (20.95, 26.08) | 21.66 (18.85, 24.76) | 21.39 (18.15, 25.02) |
| Fourth | 18.60 (16.11, 21.38) | 17.51 (15.29, 19.97) | 17.30 (14.65, 20.32) | 14.02 (11.49, 17.00) |
| Richest | 14.29 (11.99, 16.95) | 13.54 (11.58, 15.78) | 13.52 (10.73, 16.89) | 17.39 (14.09, 21.27) |
| **Sex of child** |  |  |  |  |
| Male | 52.30 (49.00, 55.58) | 50.32 (47.40, 53.24) | 53.44 (49.77, 57.08) | 49.96 (46.20, 53.71) |
| Female | 47.70 (44.42, 51.00) | 49.68 (46.76, 52.60) | 46.56 (42.92, 50.23) | 50.04 (46.29, 53.80) |
| **Maternal age** |  |  |  |  |
| 15-19 | 5.35 (4.16, 6.87) | 5.61 (4.40, 7.13) | 6.26 (4.54, 8.58) | 3.48 (2.45, 4.93) |
| 20-29 | 51.23 (47.82, 54.62) | 49.46 (46.60, 52.33) | 49.03 (45.31, 52.76) | 44.76 (40.83, 48.77) |
| 30-39 | 35.82 (32.67, 39.10) | 36.65 (33.87, 39.53) | 37.52 (33.76, 41.43) | 42.78 (38.85, 46.81) |
| 40-49 | 7.59 (6.23, 9.23) | 8.28 (6.94, 9.84) | 7.19 (5.73, 9.00) | 8.97 (7.05, 11.34) |
| **Maternal education** |  |  |  |  |
| No education | 1.66 (1.04, 2.63) | 1.58 (1.01, 2.49) | 0.79 (0.50, 1.24) | 1.15 (0.67, 1.97) |
| Primary | 22.73 (19.76, 26.01) | 19.27 (17.06, 21.68) | 14.95 (12.90, 17.26) | 11.94 (9.78, 14.50) |
| Secondary | 48.51 (45.06, 51.96) | 50.45 (47.65, 53.25) | 54.50 (50.85, 58.11) | 48.14 (44.60, 51.70) |
| Higher | 27.10 (24.28, 30.12) | 28.70 (26.11, 31.43) | 29.76 (26.17, 33.61) | 38.77 (35.07, 42.62) |
| **Place of residence** |  |  |  |  |
| Urban | 50.34 (45.24, 55.44) | 48.02 (45.66, 50.39) | 45.83 (41.43, 50.29) | 49.58 (44.27, 54.90) |
| Rural | 49.66 (44.56, 54.76) | 51.98 (49.61, 54.34) | 54.17 (49.71, 58.57) | 50.42 (45.10, 55.73) |
| **Parity** |  |  |  |  |
| One to two | 50.14 (46.81, 53.46) | 55.44 (52.62, 58.22) | 56.26 (52.38, 60.06) | 54.74 (51.04, 58.38) |
| More than two | 49.86 (46.54, 53.19) | 44.56 (41.78, 47.38) | 43.74 (39.94, 47.62) | 45.26 (41.62, 48.96) |
| **ANC visits** |  |  |  |  |
| Fewer than four | 22.54 (19.98, 25.33) | 13.67 (11.89, 15.67) | 12.82 (10.77, 15.20) | 16.18 (13.56, 19.19) |
| Four or more | 77.46 (74.67, 80.02) | 86.33 (84.33, 88.11) | 87.18 (84.80, 89.23) | 83.82 (80.81, 86.44) |
| **Distance to health facility** |  |  |  |  |
| Big problem | 28.99 (26.04, 32.13) | 27.75 (24.97, 30.71) | 25.32 (22.56, 28.29) | 16.50 (13.99, 19.37) |
| Not a big problem | 71.01 (67.87, 73.96) | 72.25 (69.29, 75.03) | 74.68 (71.71, 77.44) | 83.50 (80.63, 86.01) |
| **Maternal smoking status** |  |  |  |  |
| No | 95.23 (93.67, 96.42) | 93.61 (91.89, 94.99) | 94.43 (92.08, 96.12) | 96.25 (94.37, 97.52) |
| Yes | 4.77 (3.58, 6.33) | 6.39 (5.01, 8.11) | 5.57 (3.88, 7.92) | 3.75 (2.48, 5.63) |

### Trend in measles coverage by socio-demographic dimensions

The coverage of MCV1 showed a slight decline from 84.7% in 2008 to 80.8% in 2022. Socio-economic disparities were consistently evident across the surveys with children from the poorest wealth quintile consistently recording the lowest MCV1 coverage starting at 71.7% in 2008 and declining to 67.5% in 2022, compared to children from the richest wealth quintile with MCV1 coverage of 91.3% in 2008 and remaining stable with 91.6% in 2022. The difference in proportions in vaccination coverage between wealth quintiles across the years was statistically significant **(Supplementary Table B)**.

As with wealth quintile, other socio-demographic disparities were evident with maternal education, ANC visits, parity, and distance to health facility showing that the least advantaged groups had lower MCV1 coverage compared to the most advantaged group. Children whose mothers had secondary or higher education showed higher MCV1 coverage compared to those whose mothers had no formal education, however, we saw an increase in MCV1 coverage from 33.8% in 2008 to 54.7% in 2022 among children whose mothers had no education. Additionally, children whose mothers had four or more ANC visits consistently had higher MCV1 coverage than those children whose mothers had fewer than four ANC visits. MCV1 coverage among children whose mothers had fewer than four ANC visits declined from 68.2% in 2008 to 54.5% in 2022.

Additionally, children whose mothers had only one to two children consistently showed higher MCV1 coverage compared to those whose mothers had more than two children. We saw a decline in the MCV1 coverage among children whose mothers had more than two children from a consistent ∼80% in the first three surveys to 74.3% in 2022. Children whose household is near a health facility have higher MCV1 coverage compared to children whose household is far from a health facility. MCV1 coverage of children living far from a health facility has consistently declined from 79.4% in 2008 to 70.11% in 2022.

Mothers who don’t smoke cigarettes appear to consistently have higher MCV1 coverage for their children compared to those who smoke cigarettes. For instance, in 2022, the MCV1 coverage of children whose mothers who don’t smoke is 81.2% compared to 71.6% MCV1 coverage of children whose mothers smoke cigarettes.

MCV1 coverage according to sex of the child and age of the mother did not show substantial differences among different levels. This pattern can also be observed in urban-rural differences, except in 2017 where there appears to be a disparity, with the MCV1 coverage of children in urban areas was 85.8% compared to 77.6% for those living in rural areas. These trends are summarised in **Figure 2**.

**Figure 2:**
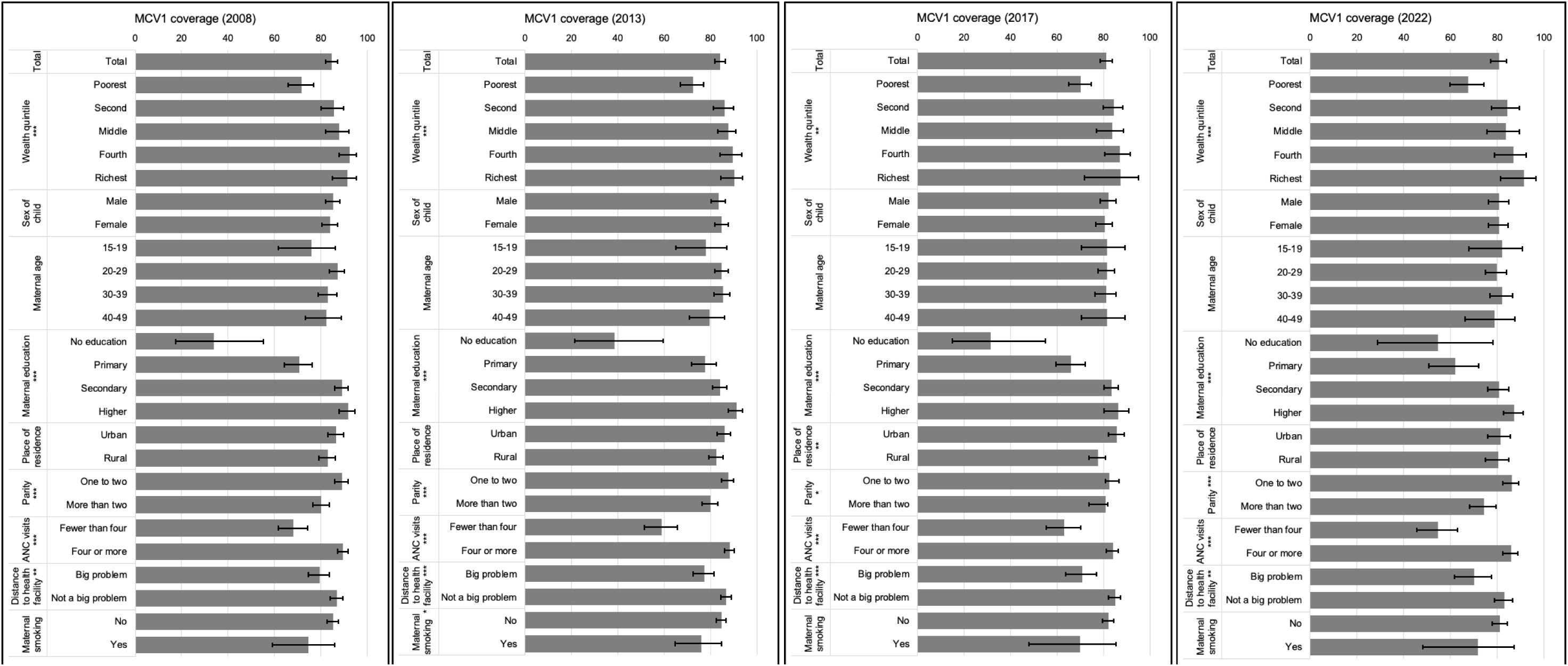
Trend in MCV1 coverage by socio-demographic characteristics of children aged 12–23 months (2008, 2013, 2017, and 2022)

### Factors associated with receiving MCV1

Using a bivariate regression analysis, we found the following independent factors to be significantly associated with receiving MCV1 dose (wealth quintile, maternal education, place of residence, parity, ANC visits, distance to health facility, and maternal smoking status). Using a multivariable logistic regression model, wealth quintile, maternal education, parity, ANC visits, distance to health facility, and maternal smoking status were all associated with receiving MCV1 dose. These factors are summarised in **Table 2** showing both the unadjusted (uOR) and adjusted Odds Ratios (aOR).

**Table 2:** Factors associated with receiving MCV1 among children aged 12-23 months (2008-2022 pooled data)

| Variables | uOR (95% CI) | p-value | aOR (95% CI) | p-value |
| --- | --- | --- | --- | --- |
| **Wealth quintile** |  |  |  |  |
| Poorest | 1.00 (ref) | ref | 1.00 (ref) | ref |
| Second | 2.42 (1.93, 3.04) | <0.001 | 1.75 (1.37, 2.25) | <0.001 |
| Middle | 2.49 (1.91, 3.24) | <0.001 | 1.51 (1.12, 2.06) | 0.008 |
| Fourth | 3.40 (2.50, 4.61) | <0.001 | 1.92 (1.32, 2.78) | 0.001 |
| Richest | 3.79 (2.36, 6.09) | <0.001 | 1.96 (1.17, 3.27) | 0.01 |
| **Maternal education** |  |  |  |  |
| No education | 1.0 (ref) | ref | 1.0 (ref) | ref |
| Primary | 3.54 (2.15, 5.83) | <0.001 | 2.24 (1.36, 3.68) | 0.002 |
| Secondary | 8.16 (5.00, 13.32) | <0.001 | 3.40 (2.05, 5.65) | <0.001 |
| Higher | 12.09 (7.2, 20.3) | <0.001 | 3.62 (2.09, 6.27) | <0.001 |
| **Place of residence** |  |  |  |  |
| Urban | 1.0 (ref) | ref | 1.0 (ref) | ref |
| Rural | 0.73 (0.60, 0.88) | 0.001 | 1.08 (0.87, 1.35) | 0.476 |
| **Parity** |  |  |  |  |
| One to two | 1.0 (ref) | ref | 1.0 (ref) | ref |
| More than two | 0.68 (0.47, 0.67) | <0.001 | 0.79 (0.65, 0.96) | 0.015 |
| **ANC visits** |  |  |  |  |
| Fewer than four | 1.0 (ref) | ref | 1.0 (ref) | ref |
| Four or more | 4.00 (3.31, 4.82) | <0.001 | 2.80 (2.31, 3.40) | <0.001 |
| **Distance to health facility** |  |  |  |  |
| Big problem | 1.0 (ref) | ref | 1.0 (ref) | ref |
| Not a big problem | 1.97 (1.61, 2.42) | <0.001 | 1.28 (1.01, 1.63) | 0.041 |
| **Maternal smoking status** |  |  |  |  |
| No | 1.0 (ref) | ref | 1.0 (ref) | ref |
| Yes | 0.55 (0.34, 0.87) | 0.011 | 0.51 (0.30, 0.87) | 0.013 |

Children from the richest wealth quintile had higher odds (aOR=1.96, 95%CI 1.17–3.27, p=0.01) of receiving MCV1 compared to children from the poorest wealth quintile. Additionally, the odds of children receiving MCV1 whose mothers had higher education (aOR=3.62, 95%CI 2.09–6.27, p<0), whose mothers had four or more ANC visits (aOR=2.80, 95%CI 2.31–3.40, p<0), and whose household is near a health facility (aOR=1.28, 95%CI 1.01–1.63, p<0.05) were higher compared children whose mothers had no education, whose mothers had less than four ANC visits, and whose household is far from a health facility.

Parity and the maternal smoking status were inversely associated with MCV1 coverage of children aged 12-23 months. Children whose mothers had more than two children had significantly lower chances (aOR=0.79, 95%CI 0.65–0.96, p<0.02) of receiving MCV1 dose compared to children whose mothers had two or less children, while children whose mothers smoke cigarettes had significantly lower odds of receiving the MCV1 dose (aOR=0.51, 95%CI 0.30–0.87, p<0.013) compared to those children whose mothers don’t smoke cigarettes.

### Trend of socio-economic inequalities in measles immunisation

The AD in predicted MCV1 coverage between the richest wealth quintile and the poorest wealth quintile appear to have slightly declined from 19.62 (95%CI 12.23–27.01) in 2008 to 17.16 (95%CI 5.16–29.15) in 2017, however it appears to have increased to 24.11 (95%CI 4.02–34.18) (as shown in **Table 3**), The SII appears to follow the same trend as shown in in **Figure 3** with an SII of 0.27 (95%CI 0.21–0.34) in 2008 that declined to 0.23 (95%CI 0.18–0.29) in 2017 and increased to 0.31 (95%CI 0.25–0.38) in 2022.

**Figure 3:**
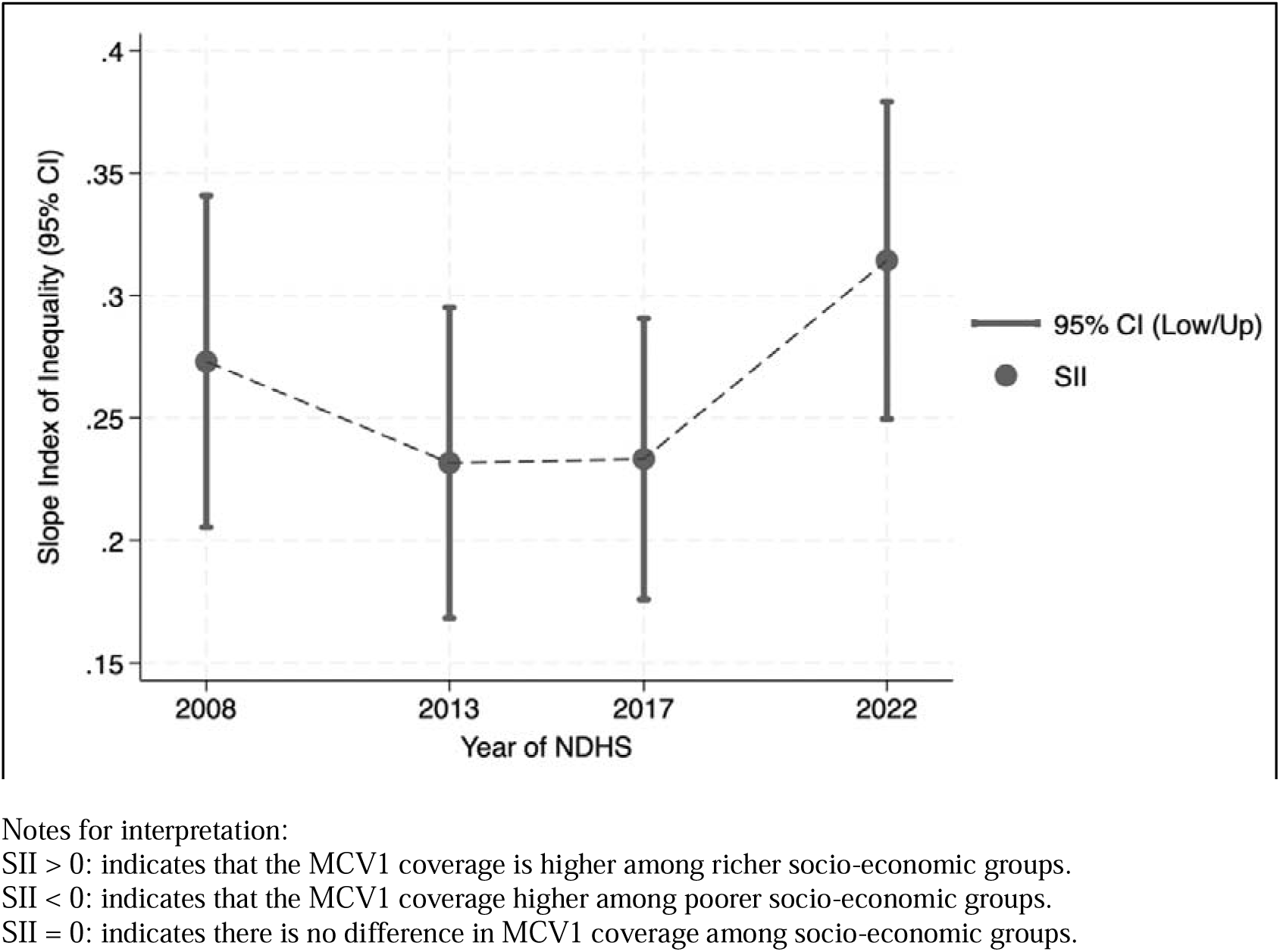
Slope Index of Inequalities (SII) of socio-economic inequalities in MCV1 coverage among children aged 12-23 months (2008, 2013, 2017, and 2022)

**Table 3:** Absolute inequalities: Crude differences and Slope Index of Inequality (SII) of socio-economic inequalities in MCV1 coverage among children aged 12-23 months (2008, 2013, 2017, and 2022)

| Absolute inequalities | 2008 | 2013 | 2017 | 2022 |
| --- | --- | --- | --- | --- |
| *Wealth quintiles (absolute difference & 95% CI)* |  |  |  |  |
| Poorest | ref | ref | ref | ref |
| Second | 13.80 (6.57, 21.03) | 13.76 (7.23, 20.29) | 14.37 (8.03, 20.70) | 16.8 (67.95, 25.77) |
| Middle | 16.14 (8.74, 23.53) | 15.24 (8.82, 21.66) | 13.49 (5.76, 21.21) | 16.06 (6.45, 25.66) |
| Fourth | 20.68 (13.96,27.39) | 17.31 (10.43,24.20) | 16.84 (9.36, 24.33) | 19.58 (9.51, 29.66) |
| Richest | 19.62 (12.23,27.01) | 17.76 (10.65,24.87) | 17.16 (5.16,29.15) | 24.11 (4.02, 34.18) |
| *Slope Index of Inequality (SII)* |  |  |  |  |
| SII (95% CI) | 0.27 (0.21, 0.34)*** | 0.23 (0.17, 0.30)*** | 0.23 (0.18, 0.29)*** | 0.31 (0.25, 0.38)*** |

p-value: (\* = p<0.05), (\*\* = p<0.01), (\*\*\*) = p<0.001)

Similarly, the relative inequalities did not show any improvement from 2008 to 2022 (as shown in **Table 4**). Across all years, the RD for the four higher wealth quintiles were consistently above 1.0, indicating inequalities experienced by the poorest wealth quintile. The RD between the richest and poorest wealth quintile have slightly declined from 1.27 (95%CI 1.19–1.33) in 2008 to 1.24 (95%CI 1.02–1.35) in 2017 but appears to have increased to 1.36 (95%CI 1.21–1.43) in 2022. The RII appears to follow the same trend as shown in **Figure 4** with an RII of 1.41 (95%CI 1.27–1.55) in 2008 that declined to 1.33 (95%CI 1.21–1.45) in 2013. This slightly increased in 2017 with an RII of 1.35 (95% CI 1.24–1.46) and then increased to 1.52 (95% CI 1.36–1.68) in 2022.

**Figure 4:**
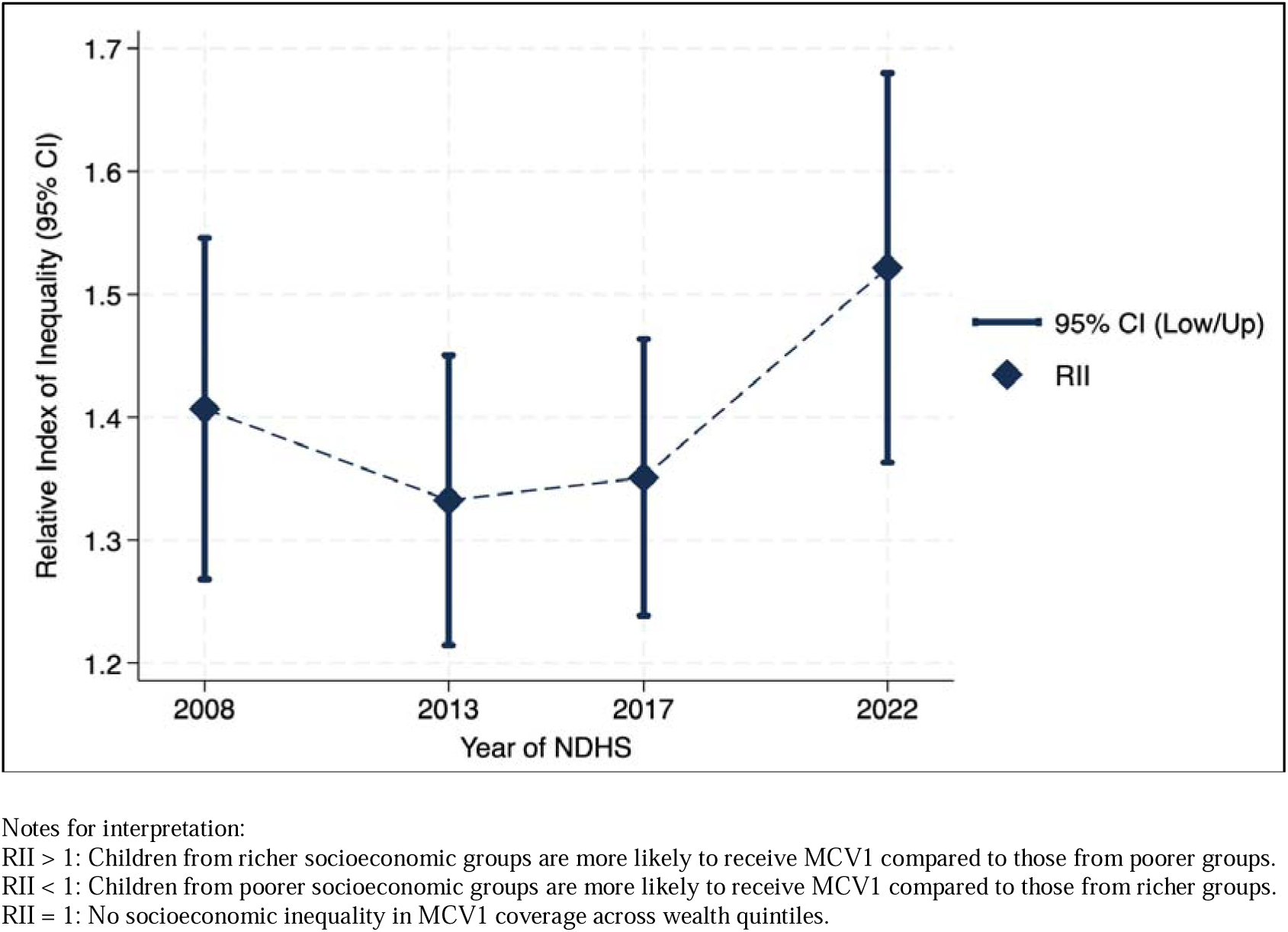
Relative Index of Inequalities (RII) of socio-economic inequalities in MCV1 coverage among children aged 12-23 months (2008, 2013, 2017, and 2022)

**Table 4:** Relative inequalities: Relative differences and Relative Index of Inequality (RII) of socio-economic inequalities in MCV1 coverage among children aged 12-23 months (2008, 2013, 2017, and 2022)

| Relative inequalities | 2008 | 2013 | 2017 | 2022 |
| --- | --- | --- | --- | --- |
| *Wealth quintiles (relative difference and 95% CI)* |  |  |  |  |
| Poorest | ref | ref | ref | ref |
| Second | 1.19 (1.12, 1.25) | 1.19 (1.12, 1.24) | 1.20 (1.14, 1.26) | 1.25 (1.15, 1.32) |
| Middle | 1.23 (1.14, 1.28) | 1.21 (1.15, 1.26) | 1.19 (1.10, 1.26) | 1.24 (1.12, 1.32) |
| Fourth | 1.29 (1.23, 1.33) | 1.24 (1.16, 1.29) | 1.24 (1.15, 1.31) | 1.29 (1.17, 1.37) |
| Richest | 1.27 (1.19, 1.33) | 1.25 (1.17, 1.30) | 1.24 (1.02, 1.35) | 1.36 (1.21, 1.43) |
| *Relative Index of Inequality (RII)* |  |  |  |  |
| RII (95% CI) | 1.41 (1.27, 1.55)*** | 1.33 (1.21, 1.45)*** | 1.35 (1.24, 1.46)*** | 1.52 (1.36, 1.68)*** |

p-value: (\* = p<0.05), (\*\* = p<0.01), (\*\*\*) = p<0.001)

Both the pooled SII [0.24, 95%CI (0.19 - 0.28), p<0.001] and pooled RII [1.75, 95%CI (1.51-2.00), p<0.001] showed socio-economic inequalities in MCV coverage.

## Discussion

We systematically analysed socio-economic inequalities in MCV1 coverage in the Philippines by identifying predictors of receiving MCV1 dose among children aged 12-23 months and examining the trends of socio-economic related inequalities in measles immunisation using both absolute and relative measures of inequalities. Our findings reveal a gradual decline in the overall national MCV1 coverage from 84.7% in 2008 to 80.8% in 2022. This downward trend coincides with a series of health system challenges [26] [27] and public health crises [28] [29] [30]. Children from the poorest wealth quintile, whose mothers had: no education; more than two children, less than four ANC visits, smoke cigarettes, and whose household is far from a health facility had lower coverage of MCV1 compared to their more advantaged respective counterparts.

In the Philippines, the persistent inadequacy in supply of measles vaccines disrupts the progress of the country in achieving the 95% coverage of life-saving vaccines [11] [26] [27]. Systemic gaps such as fragmented governance and inadequate local government funding [11], limited trained healthcare workers [26] dedicated to vaccination, and suboptimal investment vaccine infrastructure such as cold chain and logistics management [31] undermine the service delivery of measles vaccines. On the demand side, vaccine confidence in the Philippines remains fragile, influenced by multiple factors. The frequent vaccine stockouts and limited access to health facilities [11], the Dengvaxia controversy [32] which erupted in 2017, which contributed to the erosion of public’s trust to immunisation programmes, and the subsequent measles outbreak in 2019 [4] [33] and the COVID-19 pandemic in 2020 have disrupted service delivery by restricting mobility, repurposing health resources from routine to emergency, and limiting access to health facilities [3].

While national campaigns have been substantial in rebuilding measles vaccine confidence and coverage in the country, explicit equity lens in their implementation appear to be limited. A Philippine study [34] noted that communication strategies implemented by government agencies to increase childhood vaccine confidence after the Dengvaxia controversy did not reach vulnerable populations, while UNICEF Philippines [27] reported that the recovery of measles coverage has been ongoing however disparities persist with underserved areas where measles immunisation campaigns are the weakest continue to report children without any childhood vaccination. These observations underscore the limited equity lens [13] of these newly introduced interventions and recovery strategies, as they disproportionately benefit communities with existing access to vaccine information and services [13].

### Factors associated with the receipt of MCV1

Our study identified several socio-demographic factors significantly associated with MCV1 uptake among children aged 12-23 months in the Philippines. These include household wealth, maternal education, parity, ANC visits, distance to health facility, and maternal smoking status. These findings reflect broader socio-demographic inequalities in measles immunisation. Children from wealthier households (Q4 and Q5) were nearly twice as likely to receive MCV1 compared to those from the poorest wealth quintile. Maternal education was also a strong predictor, with children of mothers with secondary or higher education having over three times the odds of receiving MCV1 than those whose mothers had no formal education. These results suggest that economic stability and maternal education reduce barriers to vaccine access.

These findings are further supported by existing literature and qualitative evidence, which underscore how socio-economic status and maternal education shape access to measles immunisation services. A systematic review [35] has shown that wealth and maternal education are key determinants of access to childhood immunisation, including measles vaccines. A 2024 report [36] from the UNICEF Philippines highlights the lived experiences of mothers with low to no education who often remain unemployed and belong to lower socio-economic strata. Their children are disproportionately disadvantaged and frequently remain unvaccinated against measles due to intersecting barriers. These include unstable household income, which limits access to health services, and maternal disempowerment, which hinders mothers’ ability to understand vaccine awareness campaigns, navigate health systems and facilities, and resist misinformation about measles vaccines. [37] [38].

Geographic access also plays a critical role in measles immunisation coverage. Children from households that did not perceive distance to a health facility as a major barrier had significantly higher odds of being vaccinated (OR: 1.28 95%CI: 1.01–1.63, p=0.041) compared to those who did report it being a barrier. Despite the provision of free vaccines under Republic Act No. 10152 [10], geographic barriers continue to limit access, especially in remote and underserved regions. Public health facilities are disproportionately concentrated in economically developed northern regions, while archipelagic central areas and conflict-affected southern regions remain underserved [39] [40]. In these areas, access is further complicated by the need for boat travel [39], [41], and disruptions caused by conflict-and-violence, which damage infrastructure, interrupt cold chain systems, and displace communities [42].

ANC visits were also strongly associated with measles immunisation. Mothers who completed four or more ANC visits were significantly more likely to have their child vaccinated with MCV1(OR: 2.80, 95%CI: 2.31–3.40, p<0.001) compared to those who had less than four check-ups. ANC serves as a key entry point to maternal and child health services [43] offering opportunities for health education, vaccine awareness, and immunisation scheduling support. Regular ANC visits help reduce missed immunisation opportunities by linking mothers to services and reinforcing vaccine messaging [44], [45]. However, ANC completion is itself influenced by socio-demographic factors, with mothers with higher levels of education and better geographic access are more likely to attend the recommended number of ANC visits [46].

Parity and maternal smoking status were inversely associated with MCV1 uptake. Later-born children were less likely to be vaccinated [47], reflecting findings from previous studies that link higher parity with incomplete immunisation [48] due to time constraints, caregiver fatigue, and resource limitations [49], [50], [51]. This pattern is especially pronounced in poorer households [52] where higher parity is common despite financial constraints [53]. We found children of mothers from the richest wealth quintile (Q5) had 87.3% MCV1 coverage, compared to just 67.6% among those from the poorest wealth quintile (Q1), highlighting how socioeconomic status amplifies disparities even when household structures, such as parity, are similar.

These household-level vulnerabilities are further reflected in maternal health behaviours. Our study found that those mothers who smoke during the survey were significantly less likely to have their child vaccinated with MCV1 (OR: 0.51, 95%CI: 0.30–0.87, p=0.013) compared to those who reported they don’t smoke cigarettes. This potentially reflects broader socio-behavioural vulnerabilities and may serve as a proxy for reduced engagement with health services, lower health literacy, and diminished prioritisation of preventive care [54]. A review [54] of parental barriers to childhood immunisation found that maternal beliefs, perceptions and experiences strongly influence vaccine completion. Strengthening women’s health education and promoting health-seeking behaviours are critical in improving measles vaccine coverage and preventing future outbreak in the Philippines [55].

### Trend of socio-economic inequalities in measles immunisation in the Philippines

Our study revealed an increase in socio-economic inequalities in measles immunisation coverage in the Philippines from 2008 and 2022 as shown by the absolute and relative inequality measures. To contextualise this trend, we examined three distinct intervals, 2008–2013, 2013–2017, and 2017–2022, highlighting key policy reforms, measles-related interventions, and health crises that may have influenced the trajectory of inequality.

A modest decline in socio-economic inequalities in MCV1 was observed from 2008 to 2013 as shown by inequality measures. This coincides with equity-oriented interventions and reforms aimed at reducing gaps between the rich and the poor in accessing measles vaccines. The legislation of Republic Act No. 10152 of 2011 which institutionalised free access to childhood vaccines, including measles-containing vaccines, through public health centres nationwide [10], introduction of MCV2 to the national immunisation programmes, and strengthening of measles surveillance [56] to identify communities that are high-risk of measles transmission were implemented to further protect children from the highly infectious measles. These were aligned with the Western Pacific Regional Strategy to eliminate measles in 2012, building on the Philippines’ earlier goal of elimination by 2008 [57].

The DOH, in collaboration with WHO and UNICEF, has implemented various interventions designed to target communities that are difficult to access immunisation services. This includes SIAs conducted in 2007 and 2011 [56] aiming to vaccinated children who missed out their measles vaccine doses due. In the conflict where immunisation services have been continuously disrupted, measles vaccination strategies such as ‘Day of Peace’ [58] were launched following a ceasefire by the Moro Islamic Liberation Front. In the urban poor communities of the National Capital Region, the Reaching Every Purok (REP), an intensified reaching every community strategy by UNICEF partnered with local government units, were implemented to close the measles and polio immunisation gaps in urban slums [55] [56]. These key interventions from national to local levels highlight the success of sustained investments in ensuring high and equitable measles immunisation coverage.

Our inequality measures show stagnation in socio-economic inequalities in MCV1 coverage between 2013 to 2017, reflecting the cumulative impact of health system challenges and recurring measles outbreaks that affected the reliability and capacity of public health units to deliver routine immunisation services. Operational disruptions including measles vaccines stockouts and vaccine procurement delays have disrupted the operations of measles immunisation particularly in public health facilities located in rural and hard-to-reach areas [11], as well as in urban poor communities where overcrowding heightens the risk of transmission [59]. Further constraints in the health system such as limited number of trained healthcare workers dedicated for immunisation programmes and suboptimal cold chain infrastructure [26] [31] disproportionately impair the availability and readiness of measles immunisation programme in underserved communities [11] as wealthier families can circumvent these limitations by accessing services from private healthcare providers.

The 2013–2014 measles outbreak in the Philippines exposed persistent inequities in immunisation coverage. Despite earlier equity-focused interventions, many children from disadvantaged communities remained unvaccinated [56], [60]. The outbreak began in urban poor areas and spread to regions with weak health systems [11], underscoring fragility of local immunisation systems. During such outbreaks, routine immunisation resources such as funds, personnel, and cold chain infrastructures are diverted to emergency response such as case investigation, education campaigns, and patient management [33]. Government reports also note delays in vaccination [61], with children from wealthier households more likely to receive timely doses, while poorer families face logistical and financial barriers [62]. These findings reinforce the importance of sustained equity-focused strategies to ensure measles vaccines are consistently available and accessible in vulnerable communities.

Between 2017 and 2022, socio-economic inequalities in measles immunisation in the Philippines increased markedly. Inequality measures for 2022 surpassed the magnitudes in 2008, indicating a widening gap in measles vaccine coverage between socio-economic groups. Despite the continued measles SIAs [63], several events including a major vaccine confidence crisis, persistent operational challenges, and the COVID-19 pandemic have likely shaped this trend.

In late 2017, the Dengvaxia controversy erupted in the Philippines which centred around lack of governance and transparency, poor risk communication, and inadequate public engagement [32]. This controversy has eroded the public’s trust towards vaccines including those against measles, with a study showing that the confidence in vaccines plummeted from 93% in 2015 to just 32% in 2018 [64]. Misinformation and fear proliferated in the aftermath, disproportionately affecting low-income families who lacked access to reliable sources of information [65], [66]. This erosion of trust contributed to successive measles outbreaks, particularly in densely populated urban poor communities [29], [32]. The Dengvaxia controversy, fragmented health governance, persisting measles outbreaks, and persisting vaccine supply challenges [11] created a perfect condition to further widen the existing disparity in the receipt of measles vaccines between the rich and the poor.

The COVID-19 pandemic exacerbated these challenges. In LMICs, disruptions to childhood immunisation services led to subsequent increased mortality and outbreaks of vaccine-preventable diseases [67]. In the Philippines, movement restrictions, resource reallocation from routine to emergency response, and suspension of various immunisation campaigns among vulnerable communities [68] further limited access to measles vaccines. Loss of income and livelihood have affected the poorer households disproportionately [69], further limiting their ability to seek vaccination services in private healthcare providers. These events contributed to widening socio-economic disparities in measles immunisation and underscore the urgent need for resilient health systems and the implementation of strategies to increase measles vaccine coverage within a framework of universal health coverage and equity.

### Strengths and limitations

Our study examined trends in measles immunisation coverage by various socio-economic factors, the predictors of receiving MCV1 dose, and the trend of socio-economic inequalities using nationally representative data from four most recent DHS in the Philippines. We focused on MCV1 as an outcome as it is relevant to the Philippine setting where measles outbreaks are persistent. Measles immunisation coverage serves as an indicator of childhood immunisation programme, and the presence of measles in the community signals threat of other VPDs. This also reduced attrition and avoided reliance on other antigens. Absolute and relative inequality measures such as SII and RII were used to assess trends, and we have highlighted striking health events that might have shaped these measurements. Limitations of our study include availability of DHS data conducted before 2008, the cross-sectional analytic design used cannot establish causal relationship between explanatory variables and MCV1 status, and reliance on the information that are already collected through the DHS, with 27% of the outcome variable were based on mother’s recall and 73% on vaccination cards. Additionally, some unknown socio-demographic factors influencing measles immunisation might have been missed by the study as they are not available in the DHS.

## Conclusion

Measles is a highly infectious disease that can be prevented through safe and freely available vaccines. The Philippines has consistently experienced subnational and national outbreaks of measles, highlighting the need for high and equitable measles vaccines coverage. The country has consistently produced efforts to improve vaccine coverage to close gaps in measles vaccine and eliminate the disease; however, an overall decline in MCV1 coverage was observed while the socio-economic disparity in MCV1 surged from 2008 to 2022. These trends are likely driven by a combination of health system challenges, vaccine supply disruptions, persistent measles outbreaks, and the erosion of public trust particularly following the Dengvaxia controversy. The COVID-19 pandemic further compounded these issues, disrupting immunisation services and worsening inequalities between the rich and the poor. Despite these challenges, sustained investments in measles immunisation and strategic interventions have shown promise such as subnational policy reforms, community-based campaigns, and partnership with religious leaders that empowered local promotion of measles vaccine acceptance and access.

Although SIAs for measles are implemented to close measles vaccines inequalities, demographic barriers continue to limit vaccination among vulnerable communities [70]. The accumulation of unvaccinated children sparks pockets of measles outbreaks in the country spreading to nearby regions with suboptimal healthcare capacity, as demonstrated by the 2014, 2019, and the recent 2023-2024 measles outbreaks [7]. Monitoring of SIA-related activities from the equity lens point of view is imperative not just to increase measles vaccine coverage but also to ensure equitable access to the vaccines [13]. Finally, strengthening of primary healthcare with a social justice lens, through a robust Universal Healthcare Coverage [17], expansion of immunisation outreach activities in hard-to-reach areas, and community engagement to rebuild public trust on the measles immunisation programme are imperative in ensuring that the measles vaccines are available, accessible, and acceptable to communities regardless of their socio-demographic background.

## Data Availability

This study utilised existing datasets from the DHS website (https://dhsprogram.com). Additional access to the survey metadata was sought from the Philippine Statistics Authority (PSA).

## Acknowledgements

The authors acknowledge the technical assistance provided by the Sydney Informatics Hub, a Core Research Facility of the University of Sydney.

## Supplementary Tables

**Supplementary Table A:**
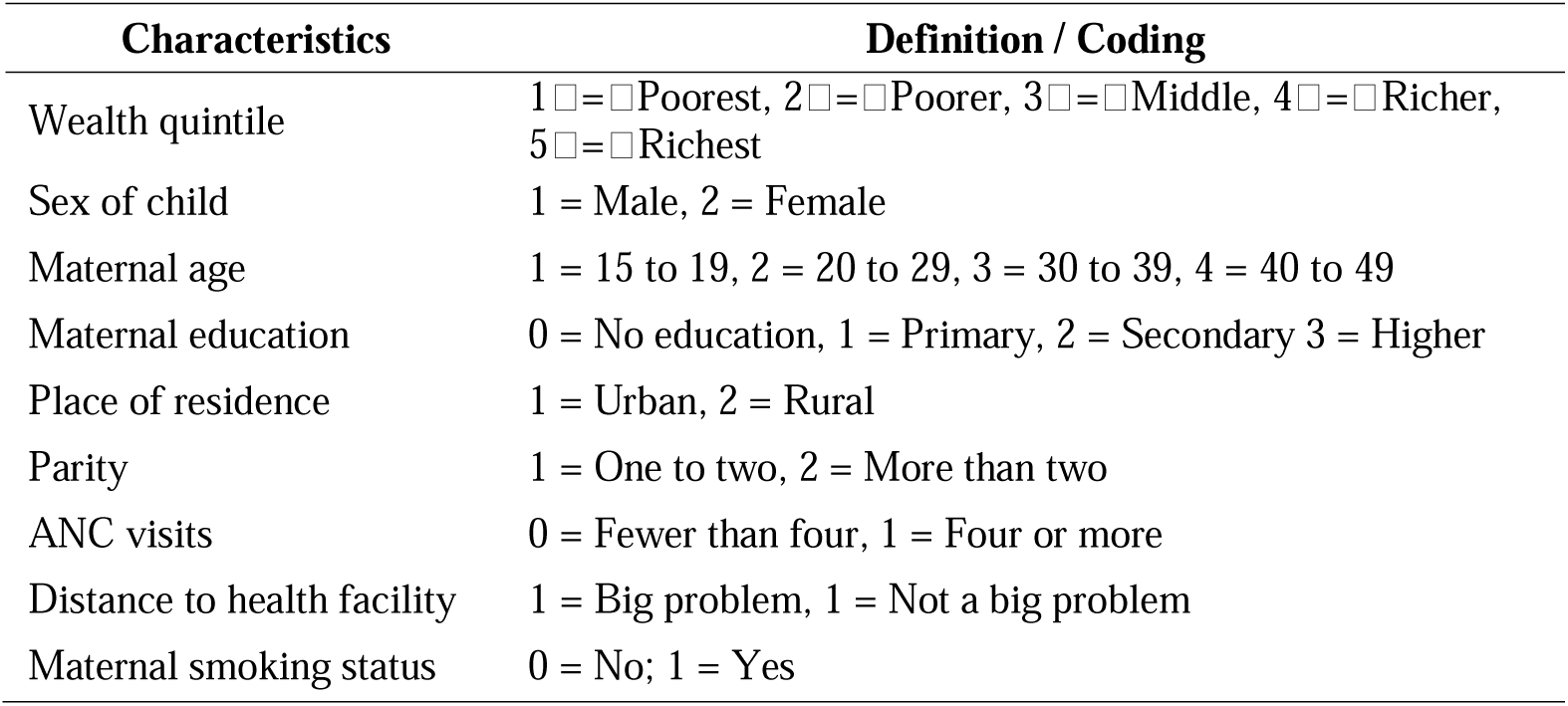
Summary of explanatory variables and their definition.

**Supplementary Table B:** Bivariate associations Between MCV1 and socio-demographic characteristics of children aged 12–23 months (2008, 2013, 2017, and 2022)

| Dimensions | 2008 N = 1163 |  | 2013 N = 1296 |  | 2017 N = 1790 |  | 2022 N = 1335 |  |
| --- | --- | --- | --- | --- | --- | --- | --- | --- |
|  | % vaccinated (95% CI) | x<sup>2</sup>-test | % vaccinated (95% CI) | x<sup>2</sup>-test | % vaccinated (95% CI) | x<sup>2</sup>-test | % vaccinated (95% CI) | x<sup>2</sup>-test |
| Total | 84.69 (82.08,86.97) |  | 84.20 (81.96,86.21) |  | 81.36 (78.75,83.71) |  | 80.81 (77.14,84.01) |  |
| Wealth quintile |  |  |  |  |  |  |  |  |
| Poorest | 71.66 (65.76,76.90) | 11.42<br>*** | 72.36 (67.09,77.08) | 10.34<br>*** | 70.20 (65.19,74.76) | 4.47** | 67.46 (59.89,74.21) | 7.17<br>*** |
| Second | 85.46 (79.91,89.68) |  | 86.12 (81.27,89.88) |  | 84.56 (79.84,88.34) |  | 84.32 (77.62,89.29) |  |
| Middle | 87.80 (81.92,91.95) |  | 87.60 (83.21,90.97) |  | 83.68 (76.92,88.75) |  | 83.51 (75.45,89.30) |  |
| Fourth | 92.34 (87.86,95.25) |  | 89.68 (84.12,93.44) |  | 87.04 (80.45,91.64) |  | 87.04 (78.71,92.43) |  |
| Richest | 91.28 (84.99,95.08) |  | 90.12 (84.33,93.93) |  | 87.35 (71.87,94.92) |  | 91.55 (81.36,96.42) |  |
| Sex of child |  |  |  |  |  |  |  |  |
| Male | 85.27 (81.90,88.11) | 0.32 | 83.59 (80.28,86.44) | 0.33 | 82.11 (78.45,85.27) | 0.44 | 80.86 (76.13,84.84) | 0.00 |
| Female | 84.04 (80.35,87.16) |  | 84.82 (81.70,87.49) |  | 80.50 (76.77,83.75) |  | 80.75 (76.04,84.72) |  |
| Maternal age |  |  |  |  |  |  |  |  |
| 15-19 | 76.00 (61.58,86.22) | 2.07 | 77.82 (64.99,86.89) | 1.37 | 81.63 (70.44,89.23) | 0.01 | 81.97 (67.80,90.75) | 0.32 |
| 20-29 | 87.06 (83.51,89.95) |  | 84.92 (81.82,87.57) |  | 81.44 (77.79,84.61) |  | 79.77 (75.02,83.81) |  |
| 30-39 | 83.09 (78.64,86.77) |  | 85.27 (81.52,88.37) |  | 81.15 (76.20,85.26) |  | 82.21 (76.99,86.45) |  |
| 40-49 | 82.29 (73.25,88.74) |  | 79.52 (70.85,86.12) |  | 81.65 (70.67,89.15) |  | 78.82 (66.35,87.54) |  |
| Maternal education |  |  |  |  |  |  |  |  |
| No education | 33.75 (17.39,55.22) | 31.71<br>*** | 38.71 (21.36,59.48) | 17.26<br>*** | 31.53 (14.83,54.92) | 14.93<br>*** | 54.68 (28.85,78.22) | 11.41<br>*** |
| Primary | 70.58 (64.18,76.26) |  | 77.69 (71.92,82.56) |  | 66.10 (59.38,72.23) |  | 61.97 (50.64,72.13) |  |
| Secondary | 89.10 (85.90,91.64) |  | 84.09 (80.83,86.89) |  | 83.49 (80.18,86.35) |  | 80.82 (75.85,84.98) |  |
| Higher | 91.74 (87.84,94.47) |  | 91.28 (87.79,93.84) |  | 86.44 (80.33,90.86) |  | 87.36 (82.70,90.90) |  |
| Place of residence |  |  |  |  |  |  |  |  |
| Urban | 86.56 (82.82,89.59) | 2.30 | 86.08 (82.95,88.70) | 2.81 | 85.83 (82.08,88.89) | 10.38** | 81.25 (76.00,85.57) | 0.06 |
| Rural | 82.79 (79.01,86.00) |  | 82.47 (79.18,85.34) |  | 77.58 (73.91,80.86) |  | 80.37 (74.98,84.83) |  |
| Parity |  |  |  |  |  |  |  |  |
| One to two | 89.16 (85.96,91.70) | 16.28<br>*** | 87.55 (84.80,89.86) | 13.05<br>*** | 82.50 (80.77,86.73) | 5.94* | 86.20 (82.47,89.24) | 15.60<br>*** |
| More than two | 80.19 (76.41,83.49) |  | 80.04 (76.46,83.21) |  | 80.87 (73.79,81.69) |  | 74.28 (68.19,79.55) |  |
| ANC visits |  |  |  |  |  |  |  |  |
| Fewer than four | 68.21 (61.62,74.14) | 66.31<br>*** | 58.84 (51.50,65.80) | 100.40<br>*** | 63.05 (55.25,70.22) | 40.02<br>*** | 54.47 (45.62,63.04) | 68.17<br>*** |
| Four or more | 89.48 (87.07,91.48) |  | 88.22 (86.08,90.07) |  | 84.05 (81.33,86.44) |  | 85.89 (82.46,88.74) |  |
| Distance to health facility |  |  |  |  |  |  |  |  |
| Big problem | 79.38 (74.46,83.56) | 9.02<br>** | 77.31 (72.46,81.53) | 16.31<br>*** | 70.77 (63.86,76.85) | 18.30<br>*** | 70.13 (61.60,77.46) | 9.62** |
| Not a big problem | 86.85 (83.85,89.37) |  | 86.85 (84.41,88.96) |  | 84.95 (82.16,87.36) |  | 82.91 (78.79,86.38) |  |
| Maternal smoking status |  |  |  |  |  |  |  |  |
| No | 85.19 (82.61,87.44) | 3.36 | 84.75 (82.56,86.71) | 3.89* | 82.03 (79.49,84.32) | 1.99 | 81.16 (77.70,84.20) | 1.20 |
| Yes | 74.70 (58.99,85.83) |  | 76.16 (64.67,84.79) |  | 69.90 (47.82,85.47) |  | 71.64 (48.24,87.25) |  |

p-value: (\* = p<0.05), (\*\* = p<0.01), (\*\*\*) = p<0.001)

